# Development of a Human Papillomavirus genotype-informed risk-stratification model to improve Cervical Cancer screening in resource-limited settings: a cross-sectional study

**DOI:** 10.64898/2026.06.06.26355059

**Authors:** Keriane Diane Kambou Kountchou, Michel Carlos Tommo Tchouaket, Larissa Gaelle Moko Fotso, Bernadette Nathalie Fokou Bomgning, Louis Fippo Fitime, Alaric Talom Teumadjou, Monique Routoube, Jeremiah Efakika Gabisa, Ezechiel Ngoufack Jagni Semengue, Alex Durand Nka, Aude Christelle Ka’e, Walter Dobgima Pisoh, Laure Deutou, Désiré Takou, Nadine Fainguem, Samuel Martin Sosso, Rachel Kamgaing Simo, Bouba Yagai, Lionel Tabola Fossa, Carlo-Federico Perno, Vittorio Colizzi, George Enow-Orock, Joseph Fokam, Alessandro Terrinoni, Jules-Roger Kuiate

## Abstract

**Background:** In resource-limited settings, a critical bottleneck in cervical cancer prevention is the lack of practical strategies to triage high-risk human papillomavirus (HR-HPV)- positive women. Therefore, this study aimed to develop and internally validate a genotype-specific risk stratification model.

**Methods:** A cross-sectional study enrolled 555 women in Cameroon. Data collection integrated cervical cytology and HPV genotyping using Abbott m2000rt and Sacace multiplex systems. An iterative modeling approach with bootstrap validation was used to develop the model and address model instability. HR-HPV genotypes were transformed into a hierarchical risk variable due to sparsity and integrated with significant predictors. The final model was translated into a scoring system, and the risk gradients and performances were evaluated at two thresholds. Data was analyzed using SPSS 27.0.

**Results:** The mean age was 44.8 years, and the prevalence of HR-HPV was 26.5% (147/555). The final model, incorporating HPV categories, age, and tobacco, demonstrated moderate discriminative ability (AUC=0.702, 0.642–0.762) with a good calibration (Hosmer-Lemeshow χ²=4.05, p=0.399). The scoring system assigned women to risk groups based on their total scores which produced a clear monotonic risk gradient; the observed probability of high-grade lesions/cancer ranged from 15% (score 0) to >65% (score ≥4). At a conservative threshold (≥4 points), 4.7% (26/555) of women were classified as high-risk, concentrating 46% (6/13) of cancers (positive predictive value[PPV]=58%) while a sensitive threshold (≥3 points) had 16.8% (93/555) high-risk, concentrating 77% (10/13) cancers (PPV=38%). Both thresholds maintained a high negative predictive value (>95%).

**Conclusion:** This bootstrap-validated, risk-stratification tool is a proof-of-concept in resource limited settings that assigns HR-HPV-positive women to distinct management pathways using three variables. After refining through a longitudinal study and external validation, this scoring system can improve the efficiency of cervical cancer screening programs in low-resource settings.

## Background

Cervical cancer remains the leading cause of cancer-related mortality among women in sub-Saharan Africa (SSA). In Cameroon, the age-standardized incidence rate is 29.6 per 100,000 women per year [1]. To address this, the World Health Organization (WHO) has set a “90–70–90” target, aiming to screen 70% of women by age 35 using high-performance molecular tests [2]. However, the transition to primary HPV screening has introduced a critical implementation bottleneck; in high-prevalence settings, the rapid increase in HPV-positive referrals often overwhelms the capacity of healthcare systems to provide colposcopy and treatment and because most HPV infections are transient and will not progress to clinically significant lesions, universal referral is neither cost-effective nor feasible in resource-limited settings [3].

Existing triage methods in Cameroon, such as Visual Inspection with Acetic Acid (VIA) and Lugol (VILI), are pragmatic but limited by; low specificity (leading to a high rate of false positives and unnecessary procedures), high variability (results are subjective and depend heavily on observer experience) [4], and limited molecular precision: binary genotyping (16/18) fails to distinguish the nuanced risk posed by diverse non-16/18 types and multiple concurrent infections. Clinical risk stratification models are essentials tools for prioritizing healthcare resources, particularly in resource limited settings where diagnostic capacity is constrained. Because referring all HPV-positive women for colposcopy and other confirmatory test (pap smear, biopsy) is not feasible nor cost effective [5], triage models that distinguish high-risk from low-risk women using molecular and readily available clinical data are essential.

A recurring challenge in building such models arises from sparse categorical predictors (variables with many possible categories, some of which are rarely observed in the sample) like HPV, with 14 or more high-risk genotypes, each representing a categorical level, with prevalence varying widely across populations [1]. Some genotypes (HPV16/18) are frequent and strongly oncogenic, while others are present and carry heterogeneous risk. Modeling each genotype as an independent binary predictor leads to instability, overfitting and unreliable coefficient estimates. Moreover, genotype risk is not binary but hierarchical, influenced by both genotype identity and the number of concurrent infections, with multiple infections conferring elevated risk [6]. This creates a multi-level predictor structure that standard modeling approaches often fail to capture efficiently.

Common strategies for managing such predictors (including regularization methods such as LASSO and category pooling) may prioritize predictive accuracy over clinical interpretability or obscure meaningful risk gradients, both of which are critical considerations for frontline implementation in resource-limited settings [7]. Transparent, parsimonious modelling strategies are therefore needed that respect the biological hierarchy of HPV risk while producing stable, interpretable coefficients.

Bootstrap resampling offers a robust framework for assessing predictor stability and model performance in such sparse data context [8]. Unlike methods that perform automatic variable selection, bootstrap validation allows to diagnose instability, guide decisions about variable aggregation based on both statistical and clinical criteria and provide optimism-corrected performance estimates [8]. This approach is particularly valuable when developing models intended for real-world clinical use, where interpretability and stability are paramount.

In this study, the primary objective was to develop and internally validate a clinically interpretable risk-stratification model for triaging HPV-positive women in resource-limited settings. Using data from our hospital-based cohort in Cameroon, we demonstrated this through: 1) diagnosing instability in individual genotype estimates via bootstrap resampling, 2) constructing a hierarchical composite HPV variable that includes genotype-specific and multiplicity-related risk, 3) developing a stable multivariate logistic regression model, 4) translating the model into an integer-based risk score with clinical decision thresholds.

## Material and Methods

### Study design and setting

We carried out a cross-sectional study that took place between March 2024 and February 2025. Participants were women who presented to the Gynecology and/or Pathology units of four hospitals across four geographic regions of Cameroon: The Bafoussam Regional Hospital (West region), the Sangmelima Reference Hospital (South region), the Garoua Regional Hospital (North region), and the Yaoundé Gynecology, Obstetric and Pediatric Hospital (Centre region).

### Participant

We included sexually active women aged 25 years and older who visited the pathology and or gynecology units of participating hospitals during the study period and provided written informed consent (Fig 1). We excluded pregnant women, those who had undergone cervical excision or hysterectomy and those who were undergoing any cervical therapy for cervical cancer. A convenient sampling method was used whereby all eligible women who consented were enrolled. Sociodemographic characteristics of the study population were broadly compared with national demographic data from Cameroon. Ethical approval was obtained from the National Committee for Ethics in Human Health Research (No. 2020/06/1249/CE/CNERSH/SP) and institutional authorizations for laboratory analyses and sample collection were obtained from each participating site.

**Fig 1:**
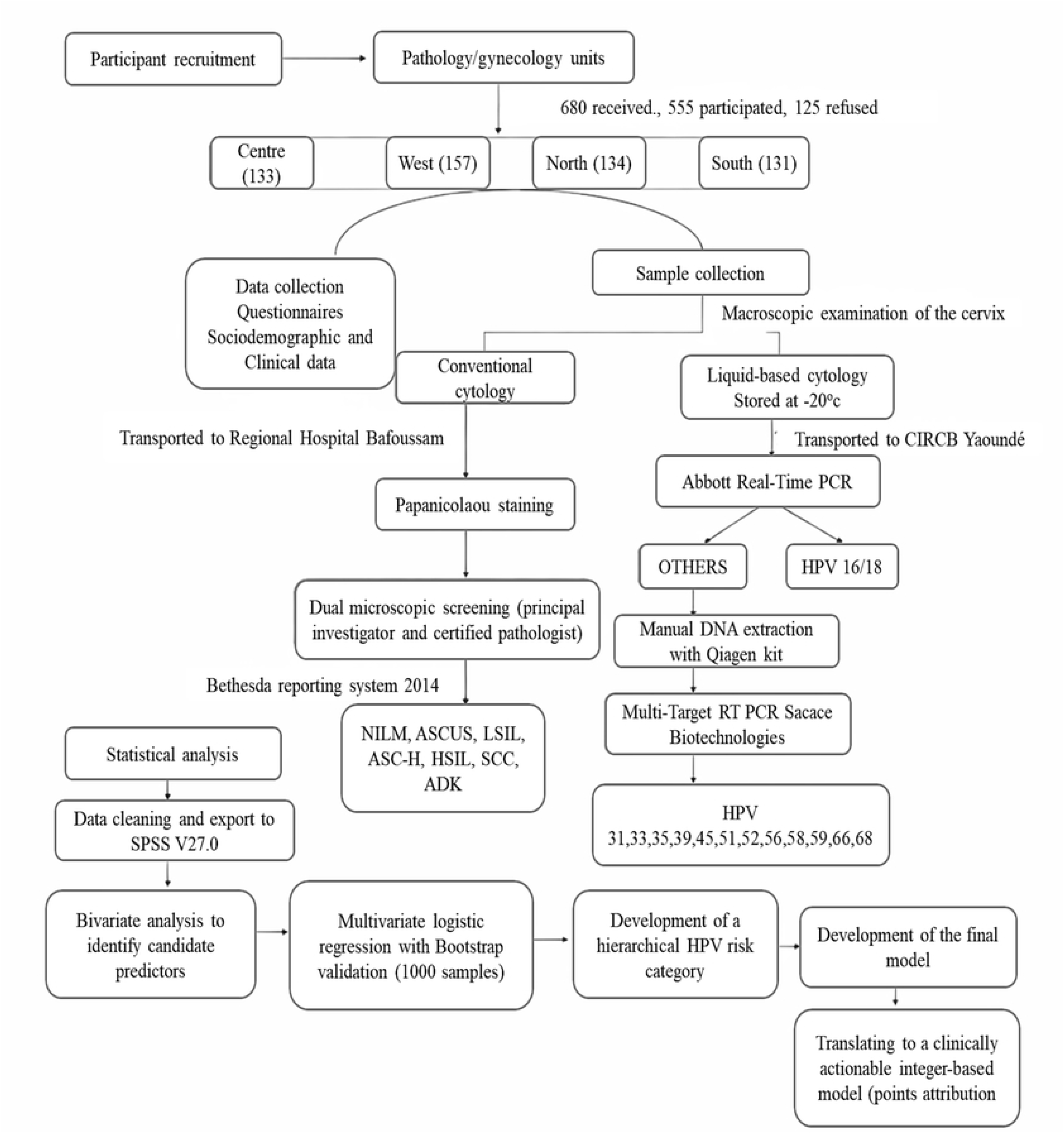
Flowchart of participant recruitment, sample collection, laboratory procedures, and statistical analysis.

Of 773 women assessed for eligibility, 93 were excluded (34 pregnant, 21 with prior hysterectomy, 38 undergoing cervical cancer treatment), leaving 680 eligible. Of these, 125 declined participations, resulting in a final analytic sample of 555 participants.

### Data preparation and quality checking

All laboratory procedures included internal quality controls. Cytology slides with ambiguous findings underwent blinded dual review by two independent pathologists. HR-HPV genotype detection followed standardized protocols on validated platforms (Abbott m2000rt and Sacace Biotechnologies).

Post-hoc calculations for sample size indicated there were 81 patients with high grade lesions and cancer (≥HSIL) in the final dataset. The EPV calculation was based on a preliminary dataset of 81 events yielding EPV of 27 which is well above the minimum recommended threshold of 10 to 20 for stable multivariable logistic regression analysis [9].

### Outcome definition

A strict distinction was maintained between the outcomes used. The primary outcome was ≥HSIL, defined as the presence of a high-grade squamous intraepithelial lesion or worse, comprising high-grade squamous intraepithelial lesions (HSIL), squamous cell carcinoma (SCC), or adenocarcinoma (ADK) on cytology, according to the Bethesda System 2014. A broader secondary outcome (≥LSIL, defined as any cytological abnormality other than NILM) was used exclusively in exploratory bivariate analyses to identify candidate predictors and was not included as a performance metric in any other analysis.

### Predictor variables

Predictor selection was based on clinical relevance and supporting evidence in the literature. Candidate predictors comprised individual HR-HPV genotypes (types 16, 18, 31, 33, 35, 39, 45, 51, 52, 56, 58, 59, 66, and 68), the total number of concurrent HR-HPV infections, and clinical/demographic variables including age, parity, marital status, education level, occupation, tobacco use, alcohol consumption, contraceptive use, HIV status, number of lifetime sexual partners, and age at first sexual intercourse. None of the predictors was selected by automated means. An age cut-off of 36 years was derived from receiver operating characteristic (ROC) analysis optimizing sensitivity and specificity for ≥HSIL detection, consistent with epidemiological evidence of higher HPV persistence risk beyond age 35 in sub-Saharan Africa.

### Data collection and laboratory procedures

Cervical samples were collected from all participants for both liquid-based and conventional (Pap smear) cytology. Pap smears were stained using Papanicolaou staining method and reported according to the Bethesda System 2014.

HR-HPV genotyping was performed using Abbott Real Time HR-HPV assay (m2000 system), which detected 14 HR-HPV genotypes with separate identification of HPV 16 and 18 and pooled detection of 12 other high-risk genotypes. Samples positive for the later further underwent extended genotyping through manual DNA extraction (Qiagen QIAamp DNA mini kit) followed by multiplex real-time PCR (Sacace Biotechnologies HPV genotype 14 Real-TM Quant System).

### Missing data

There was no missing data for either HR-HPV predictor variables or for the primary outcome. A small percentage of missing data (<2%; n = 9) came from the questionnaire-derived covariates and given their low percentage and random distribution, complete-case analysis was deemed suitable for the evaluation.

### Reference standards

Although histopathology is considered the gold standard, its systematic use in large-scale studies in low-resource settings is limited by ethical, logistical, and financial constraints. In this study, cytology was used as the highest routinely available diagnostic standard. To ensure reliability, all slides were independently reviewed by two experienced cytopathologists, with discrepancies resolved by consensus, in accordance with the Bethesda 2014 system.

### Statistical analysis and model development

Statistical analyses were established by using SPSS (IBM Corporation) Version 27.0. The modeling process consisted of eight sequential steps:

1. Exploratory analysis: Bivariate logistic regression analyses were conducted using the broader outcome (≥LSIL) to identify predictors associated with cervical lesions to be used in model development. Variables with p<0.05 were retained for multivariate logistic regression analysis.
2. Assessment of HPV genotype instability: An initial multivariate logistic regression model including all 14 HR-HPV genotypes, with ≥HSIL as outcome was done. Model fit was assessed using the likelihood ratio chi-square test and explained variance using Nagelkerke’s R^2^. Internal validation was performed using bootstrap resampling (1000 replicates). Bias corrected and accelerated (BCa) 95% confidence intervals were estimated for each genotype ratio to assess coefficient stability. Genotypes exhibiting extreme instability, wide confidence intervals spanning null, or inconsistent effects were considered unsuitable for the next modeling stage.
3. Intermediate multivariate model: An intermediate model incorporating stable genotypes and retained clinical covariates was fitted using ≥HSIL as the outcome. Bootstrap validation (1000 replicates) was applied to evaluate predictor stability; variables with unstable estimates or implausibly wide confidence intervals were excluded from further modelling.
4. Construction of a hierarchical HPV risk variable: To address data sparsity and capture both genotype-specific and multiplicity-related risk, a five-level hierarchical HPV risk variable was constructed based on statistical evidence and biological plausibility as follows; HPV-negative (reference), single non-16 HR-HPV infection, multiple non-16 HR-HPV infections, HPV 16 alone, and HPV 16 with one or more additional genotype(s). This helped reduce dimensionality while preserving a clinically meaningful risk gradient.
5. Final model development and internal validation: The final multivariate logistic regression model incorporated the hierarchical HPV risk categories and stable clinical predictors identified in preceding steps. Model selection was guided by clinical relevance, statistical significance (p<0.05) and bootstrap stability. Internal validation was conducted using 1000 bootstrap resamples to estimate optimism-corrected model performance and assess coefficient robustness. Firth’s penalized likelihood regression was applied to statistically significant and biologically plausible predictors with very wide confidence intervals to correct for quasi-complete separation.
6. Model performance assessment: Discrimination was evaluated using the area under the receiver operating characteristic curve (AUC) with 95% confidence intervals. Calibration was assessed using a calibration plot and the Hosmer-Lemeshow goodness of fit test comparing of observed versus predicted risk across risk deciles.
7. Development of the integer risk score: To facilitate clinical implementation, odds ratios from the final model were converted into an integer-based risk score. Points were assigned using logarithmic scaling of adjusted odds ratios as follows; OR 2.0 – 3.9 = 1point, OR 4.0 – 7.9 = 2points, OR 8.0 – 15.9 = 3points and OR ≥16 = 4points. Individual risk scores were calculated by summing points across all predictors.
8. Evaluation of high-risk thresholds: The risk scoring system was then applied to our study population (555 women), and the risk gradient was evaluated by examining the proportion of ≥HSIL across score categories. Two implementation high-risk thresholds were evaluated: a conservative threshold (≥4 points) to optimize resource use and a sensitive threshold (≥3 points) to maximize cancer detection. These thresholds were chosen because there was no statistically significant difference between thresholds ≥3 and ≥4 points (p=0.16) compared to threshold ≥2 and ≥3 (p<0.001) and ≥2 and ≥4 (p<0.0001). Sensitivity, specificity, positive predictive value (PPV) and negative predictive value (NPV) were calculated at each threshold. Specificity was calculated as True Negative (TN)/TN + False Positive (FP), where TN represented women without ≥HSIL correctly classified as low risk and FP represented women without ≥HSIL incorrectly classified as high risk.

## Results

### Sociodemographic characteristics and baseline data

A total of 555 women were enrolled in the study after consenting to participate. The mean age was 44.8 years (±13.2) with the majority having secondary (44.1%) or university (33.5%) education. Most participants were married (56.8%), and housewives represented the largest occupational group (31.7%).

The overall prevalence of HR-HPV was 26.5 (147/555) with the most prevalent genotype among HPV-positive women being HPV 16 (21.1%, 31/147). High grade cervical precancerous lesions and cancer (HSIL+) represented 14.6% (81/555). Baseline sociodemographic and clinical characteristics derived from robust and validated laboratory procedures (cytology and HPV genotyping) are presented in Table 1.

**Table 1:** Baseline sociodemographic and clinical characteristics of the study population (N=555)

| Variable | Category | n (%) |
| --- | --- | --- |
| Age (years) † | Mean ± SD | 44.8 ± 13.15 |
|  | 25–35 | 148/555 (26.7%) |
|  | 36–57 | 319/555 (57.4%) |
|  | >57 | 88/555 (15.9%) |
| HR-HPV status † | Positive | 147/555 (26.5%) |
| Most prevalent HR-HPV type (among HPV+) | HPV 16 | 31/147 (21.1%) |
| HIV status † | Positive | 76/555 (13.7%) |
| Cytology (Bethesda 2014) | NILM | 373/555 (67.2%) |
|  | ≥LSIL (outcome for bivariate analysis) | 182/555 (32.8%) |
|  | ≥HSIL (primary outcome) | 81/555 (14.6%) |
|  | SCC | 13/555 (2.3%) |
|  | ADK | 2/555 (0.4%) |
| Tobacco use † | Yes | 53/555 (9.5%) |
| Lifetime sexual partners | 1–2 | 132/555 (23.8%) |
|  | ≥3 | 423/555 (76.2%) |
| Parity † | 0 | 54/555 (9.7%) |
|  | 1–3 | 191/555 (34.4%) |
|  | ≥4 | 310/555 (55.9%) |
| Contraceptive use | Yes | 219/555 (39.5%) |
| Alcohol use † | Yes | 338/555 (60.9%) |

*Note: NILM: negative for intraepithelial lesion or malignancy; ASC-US: atypical* *squamous cells of undetermined significance; LSIL: low-grade squamous intraepithelial* *lesion; ASC-H: atypical squamous cells, cannot exclude HSIL; HSIL: high-grade* *squamous intraepithelial lesion; SCC: squamous cell carcinoma; ADK:* *adenocarcinoma. † p<0.05 in bivariate analysis with ≥LSIL outcome.*

### Selection of candidate variables

Using the broader cervical lesions (≥LSIL) as outcome, bivariate logistic regression analyses identified several candidate predictors for further modeling. HR-HPV, number of concurrent HPV infections, age, parity, HIV-positive status, tobacco use and alcohol consumption showed significant associations (p<0.05) and were retained for model development. Number of sexual partners, age at first sexual intercourse, contraceptive use, sexually transmittable diseases, marital status and educational level (p>0.05) were not statistically significant and were excluded [S1 Table]

### Model development and validation (multivariate logistic regression with bootstrap resampling)

The first multivariate model containing all 14 HPV genotypes with ≥HSIL (high grade cervical lesions and cancer) as outcome showed significant overall fit (χ²=37.22, df=14, p=0.001), explained limited variance (Nagelkerke R²=0.12) and bootstrap validation (1000 replicates) revealed substantial instability for most genotypes (Table 2, S1 Fig). Only HPV 16 demonstrated both statistical significance and bootstrap stability (OR=6.97, 95% CI: 2.47–22.11, p<0.001). Other genotypes showed wide CIs spanning null values, confirming their unsuitability for individual modeling in this dataset.

**Table 2:** Logistic regression analysis of individual HPV genotypes as predictors of cervical lesions, with bootstrap validation (n=1000 samples)

| HPV subtype | N (%) | Adjusted OR (95% CI) | p-value | Conclusion |
| --- | --- | --- | --- | --- |
| 16 | 31 (21.09) | 6.97 (2.47 – 22.11) * | <0.001 | Robust effect |
| 18 | 13 (8.84) | 0.23 (0.00 – 0.82) * | 0.241 | Unstable, wide CI |
| 31 | 17 (11.56) | 2.47 (0.37 – 12.45) * | 0.151 | Not significant |
| 33 | 17 (11.56) | 1.60 (0.45 – 3.55) * | 0.460 | Not significant |
| 35 | 18 (12.24) | 1.64 (0.13 – 7.05) * | 0.441 | Not significant |
| 39 | 7 (4.76) | 0.88 (0.00 – 8.98) * | 0.911 | Extreme instability |
| 45 | 22 (14.97) | 3.09 (0.37 – 60.66) * | 0.056 | Borderline, imprecise |
| 51 | 10 (6.80) | 1.45 (0.00 – 6.49) * | 0.668 | Unstable |
| 52 | 15 (10.20) | 1.93 (0.24 – 8.41) * | 0.331 | Not significant |
| 56 | 19 (12.93) | 1.13 (0.21 – 3.87) * | 0.849 | Not significant |
| 58 | 21 (14.29) | 1.70 (0.40 – 4.61) * | 0.371 | Not significant |
| 59 | 7 (4.76) | 2.89 (0.00 – 213M) * | 0.261 | Extreme instability |
| 66 | 17 (11.56) | 1.38 (0.05 – 7.72) * | 0.643 | Not significant |
| 69 | 28 (19.05) | 0.41 (0.07 – 1.04) * | 0.197 | Not significant |

*Reference category: HPV negative. \* = Bootstrap Bias Corrected Accelerated (Bca) 95%* *CI from 997 samples. Model $X^2 = 37.22$ , $df = 14$ , $p=0.001$ , Nagelkerke $R^2 = 0.12$*

The instability was mitigated when we replaced all the genotype predictor variables with a five-level hierarchical HPV variable, ultimately yielding stable, clinically meaningful estimates while eliminating quasi-complete separation. After iterative selection retaining only predictors with stable bootstrap-corrected estimates and established biological significance [S2 Table], the final model contained three predictors: hierarchical HPV risk category, age ≥36 years, and tobacco use (Table 3).

**Table 3:** Final multivariable logistic regression model (N=555; outcome: ≥HSIL). Bootstrap-corrected AUC = 0.702 (95% CI: 0.64–0.76)

| Predictors | Categories | Adjusted OR (95% CI) | p-value |
| --- | --- | --- | --- |
| HPV risk categories | HPV 16 alone Vs negative | 7.73 (2.75 – 22.35)<sup>b</sup> | < 0.001 |
|  | Single non-16 Vs negative | 1.75 (0.74 – 3.62)<sup>b</sup> | 0.143 |
|  | Multiple non-16 Vs negative | 2.71 (1.11 – 5.63)<sup>b</sup> | 0.014 |
|  | 16 + coinfections Vs negative | 28.67 (2.53 – 58.6M)<sup>b</sup> | < 0.001 |
|  | 16 + coinfections firth-corrected | 12.40 (3.10 – 49.80) | < 0.001 |
| Age (years) | [36 – 57] Vs [25 – 35] | 4.69 (1.81 – 46.45)<sup>b</sup> | < 0.001 |
|  | > 58 Vs [25 – 35] | 6.69 (2.25 – 60.11)<sup>b</sup> | < 0.001 |
| Tobacco use | YES Vs NO | 3.04 (1.40 – 6.30)<sup>b</sup> | 0.002 |

*Note: Reference categories: HPV negative, age [25 – 35] years, tobacco non-consumption and HIV negative. <sup>b</sup> = Bootstrap Bca 95% CI from 999 samples. Model $X^2 = 59.0$ , $df = 8$ , $p < 0.001$ , Nagelkerke $R^2 = 0.179$ . Firth's penalised likelihood regression was applied to the HPV 16 + co-infections category to correct for quasi-complete separation. Model discrimination was moderate (bootstrap-corrected AUC = 0.702, 95% CI: 0.64–0.76), and calibration was acceptable (Hosmer–Lemeshow $\chi^2 = 4.05$ , $df = 4$ , $p = 0.399$ ).*

The model demonstrated a clear risk gradient with bootstrap validation (1000 replicates): compared with HPV-negative women, those with single non-16 infections showed a non-significant increase in odds (OR = 1.75, p = 0.143), multiple non-16 infections conferred a 2.71-fold increase (95% CI: 1.11–5.63, p = 0.014), HPV 16 alone conferred a 7.73-fold increase (95% CI: 2.75–22.35, p < 0.001), and HPV 16 with co-infections showed markedly elevated odds (asymptotic OR = 28.67; Firth-corrected OR = 12.40, 95% CI: 3.10–49.80, p < 0.001). Age and tobacco use retained strong, stable associations.

### Model discrimination and calibration

Internal bootstrap validation (1000 replicates) yielded an optimism-corrected AUC of 0.702 (95% CI: 0.64–0.76), consistent with internationally accepted benchmarks for cervical cancer screening triage tools and indicative of moderate discriminative ability [10] (Fig 2).

**Fig 2:**
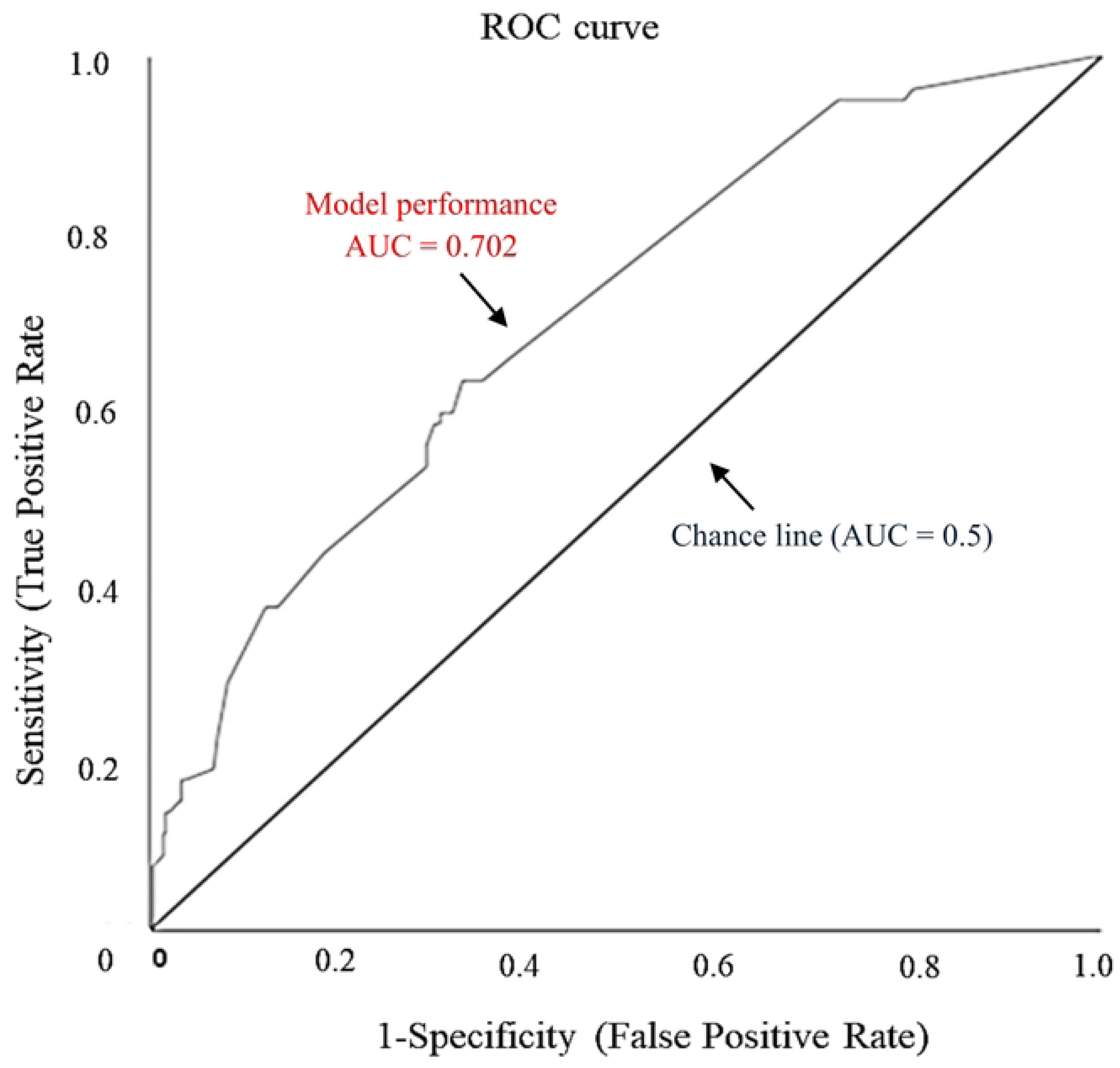
Receiver Operating Characteristic (ROC) curve for the final risk-stratification model.

There was no evidence of poor calibration (Hosmer–Lemeshow χ² = 4.05, df = 4, p = 0.399); the calibration plot showed good agreement between predicted probabilities and observed ≥HSIL rates across risk deciles, with a near-zero observed event rate in the lowest-risk deciles. The diagonal represents perfect calibration, and the dots indicate observed probabilities across deciles (Fig 3).

**Fig 3:**
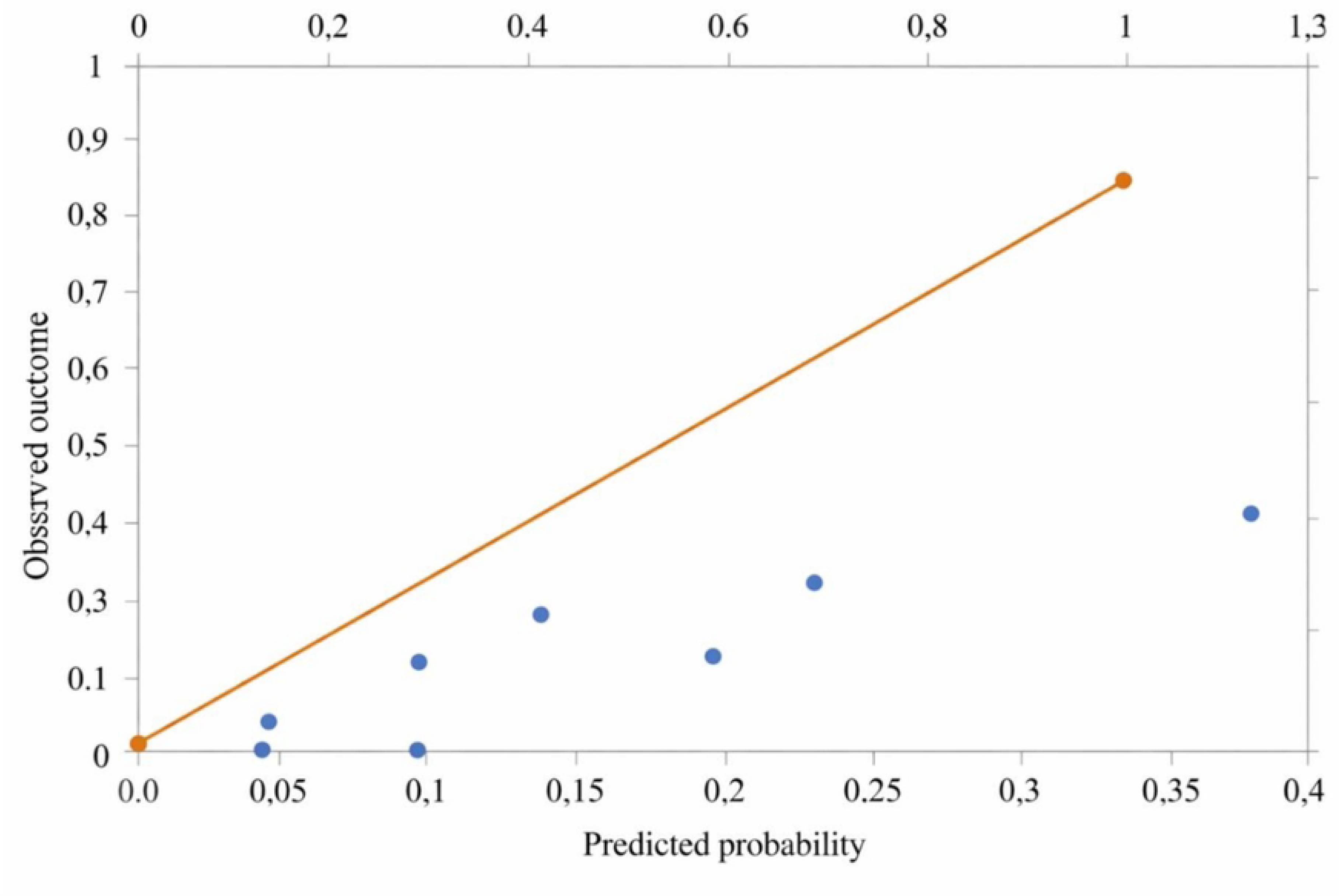
Calibration plot for ≥HSIL prediction.

### Integer risk scoring system

The predictors’ odds ratios from the final model were converted to a practical integer scoring system to facilitate clinical implementation (Table 4). The number of points assigned was proportional to these adjusted odds ratios using a logarithmic scale. The maximum score possible is 6 points (HPV 16 + co-infection [3 pts] + 36 years of age or older [2 pts] + tobacco use [1 pt]).

**Table 4:** Illustration of the scoring system of predictor variables based on their odd ratios.

| Predictors | Categories | Adjusted OR | Score |
| --- | --- | --- | --- |
| High-risk HPV | HPV negative (reference) | 1 | 0 |
|  | HPV 16 alone | 7.73 | 2 |
|  | Single non 16 | 1.75 | 0 |
|  | Multiple non 16 | 2.71 | 1 |
|  | HPV 16 + coinfections* | 12.40 | 3 |
| Age (years) | [25 – 35] (reference) | 1 | 0 |
|  | [36 – 57] | 4.69 | 2 |
|  | >58 | 6.69 | 2 |
| Tobacco consumption | No (reference) | 1 | 0 |
|  | Yes | 3.04 | 1 |
| Maximum possible score |  |  | 6 |

*Note: \* OR shown is the asymptotic estimate from the initial model (OR = 28.674), retained for scoring purposes due to its position in the logarithmic scale ( $\geq 16 = 4$ pts). This estimate is statistically unstable due to quasi-complete separation. The Firth-corrected OR = 12.40 (95% CI: 3.10–49.80) is the preferred estimate for interpretation (see Table 3). Scoring rule: OR 2.0–3.9 = 1 pt; 4.0–7.9 = 2 pts; 8.0–15.9 = 3 pts; $\geq 16.0 = 4$ pts.*

### Risk gradient and threshold evaluation

The final combined risk score yielded a strong monotonic gradient of risk for ≥HSIL at all score levels (Fig 4): The probability of having an ≥HSIL increased from about 15% at a score of 0 to 33% at a score of 2, to 49% at score level 3, and to ≥65% at score levels ≥4. This graded relationship confirms the scoring system’s ability to stratify women into distinct risk categories. Performance characteristics at the two pre-specified clinical high-risk thresholds are summarized in Table 5.

**Fig 4:**
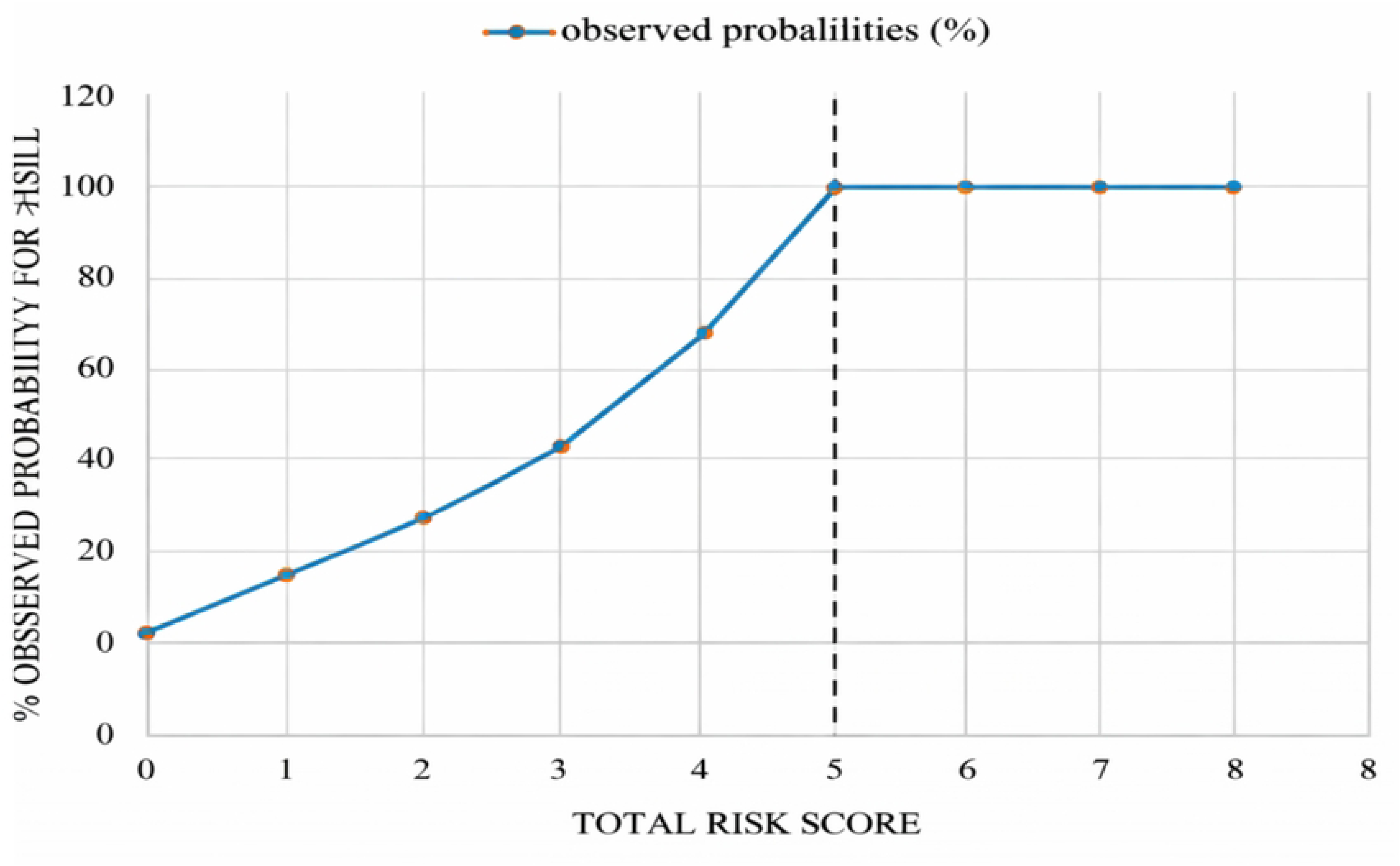
Observed probability of ≥HSIL by total risk score (monotonic risk gradient)

**Table 5:** Diagnostic performance of the scoring system at conservative (score ≥4) and sensitive (score ≥3) thresholds (primary outcome: ≥HSIL)

| Performance metric | Conservative (Score $\geq 4$) | Score $\geq 4$ 95% CI | Sensitive (Score $\geq 3$) | Score $\geq 3$ 95% CI |
| --- | --- | --- | --- | --- |
| Women classified as high-risk, n (%) | 26 (4.7%) | — | 93 (16.8%) | — |
| Sensitivity | 18.5% | 0.12 – 0.28 | 43.2% | 0.33 – 0.54 |
| Specificity | 27.2% | 0.23 – 0.32 | 23.2% | 0.20 – 0.27 |
| PPV | 57.7% | 0.39 – 0.75 | 37.6% | 0.28 – 0.48 |
| NPV | 96.3% | 0.92 – 0.99 | 95.7% | 0.90 – 0.98 |
| Cancers captured, n/total (%) | 6/13 (46%) | — | 10/13 (77%) | — |
| Referral rate | 4.7% | — | 16.8% | — |

*PPV: positive predictive value; NPV: negative predictive value. Primary outcome: ≥ HSIL (HSIL, SCC, ADK). Referral rate = proportion of total cohort classified as high-risk.*

The risk scoring system was then applied to our study population (555 women) and risk score distributions and corresponding risk metrics by stratum are presented in Tables 6a and 6b. When a conservative threshold (≥4 points) was applied to define high risk (Table 6a), only 4.7% (n=26/555) of the cohort was classified as high risk, which concentrated 73.1% (19/26) of lesions (≥LSIL) and 46% (6/13) of cancers (SCC). The risk of having any cervical lesion (≥LSIL) was observed in 73.1% of women, and 23.1% (6/26) had invasive SCC. Compared to the low-risk group (scores 0 – 1), women in the high-risk group had a 4.5-fold increased relative risk (RR=4.46, 95% CI: 2.96 – 6.72) for any cervical lesion. A clear stepwise risk gradient was observed across strata: low → moderate (2.2-fold increase) and moderate → high (2.0-fold increase).

**Table 6a:** Risk score distribution and stratum-specific risk metrics: conservative threshold (score ≥4)

| Score | n (%) | Observed ≥LSIL risk (%) | SCC prevalence (%) | Relative risk (95% CI) | Risk category |
| --- | --- | --- | --- | --- | --- |
| 0–1 | 134 (24.1) | 16.4 | 0 | Reference | Low |
| 2–3 | 395 (71.2) | 35.7 | 1.8 | 2.18 (1.46–3.25) | Moderate |
| $\geq 4$ | 26 (4.7) | 73.1 | 23.1 | 4.46 (2.96–6.72) | High |

*Observed risk = prevalence of any cytological abnormality ( ≥ LSIL) within each stratum. Relative risk vs. low-risk reference group (score 0–1). Risk gradient: Low → Moderate = 2.2-fold; Moderate → High = 2.0-fold.*

**Table 6b:** Risk score distribution and stratum-specific risk metrics: sensitive threshold (score ≥3)

| Score | n (%) | Observed ≥LSIL risk (%) | SCC prevalence (%) | Relative risk (95% CI) | Risk category |
| --- | --- | --- | --- | --- | --- |
| 0 | 115 (20.7) | 14.8 | 0 | Reference | Low |
| 1–2 | 347 (62.5) | 32.9 | 0.9 | 2.22 (1.40–3.54) | Moderate |
| $\geq 3$ | 93 (16.8) | 53.8 | 10.8 | 3.64 (2.30–5.76) | High |

*Observed risk = prevalence of any cytological abnormality ( ≥ LSIL) within each stratum. Relative risk vs. low-risk reference group (score 0). Risk gradient: Low → Moderate = 2.2-fold; Moderate → High = 1.6-fold.*

To improve case detection, a sensitive threshold (≥3 points) was evaluated, classifying 16.8% (n=93/555) as high risk (Table 6b). This group exhibited 3.6-fold increased risk (RR=3.64, 95% CI: 2.30–5.76) compared to low-risk group, concentrating 54.8% (51/93) of lesions and capturing 77% (10/13) of all SCC cases. The risk gradient from moderate to high was 1.6-fold, reflecting the trade-off between broader referrals.

Cytological distribution across risk categories further validated the gradient (Table 7).

**Table 7:** Distribution of cervical cytology findings by risk stratum (conservative threshold: score ≥4)

| Cytology | Low risk (score 0–1) n=134 | Moderate risk (score 2–3) n=395 | High risk (score $\geq 4$) n=26 | Total N=555 |
| --- | --- | --- | --- | --- |
| NILM | 112 (83.6%) | 254 (64.3%) | 7 (26.9%) | 373 (67.2%) |
| ASC-US | 3 (2.2%) | 11 (2.8%) | 1 (3.8%) | 15 (2.7%) |
| LSIL | 13 (9.7%) | 59 (14.9%) | 3 (11.5%) | 75 (13.5%) |
| ASC-H | 1 (0.7%) | 8 (2.0%) | 0 (0%) | 9 (1.6%) |
| HSIL | 5 (3.7%) | 54 (13.7%) | 9 (34.6%) | 68 (12.3%) |
| SCC | 0 (0%) | 7 (1.8%) | 6 (23.1%) | 13 (2.3%) |
| ADK | 0 (0%) | 2 (0.5%) | 0 (0%) | 2 (0.4%) |
| Total | 134 (24.1%) | 395 (71.2%) | 26 (4.7%) | 555 |

$\chi^2(12, N=555) = 87.34, p < 0.001$ , Cramér's $V = 0.27$ . key findings: 0% SCC in the low-risk group; 23.1% SCC in the high-risk group. NILM: negative for intraepithelial lesion or malignancy.

### Clinical implementation algorithm

A two-threshold clinical algorithm was created to help translate the scoring system to front-line management decision making (Fig 5). In settings with severe resource constraints, the conservative threshold (≥4 points) is recommended: it targets only 4.7% of women at highest risk (PPV = 57.7%), thereby maximizing resource efficiency. In programs prioritizing cancer detection, the sensitive threshold (≥3 points) is recommended: it broadens referrals to capture 77% of cancers while maintaining a moderate PPV (37.6%). Clinical recommendations include women in the low-risk group (0–1 points) should undergo repeat screening after one year; those in the moderate-risk group (2–3 points) warrant six-month follow-up with cytology (Pap smear); women in the high-risk group should be referred promptly for colposcopy, biopsy, or ablative treatment as indicated.

**Fig 5:**
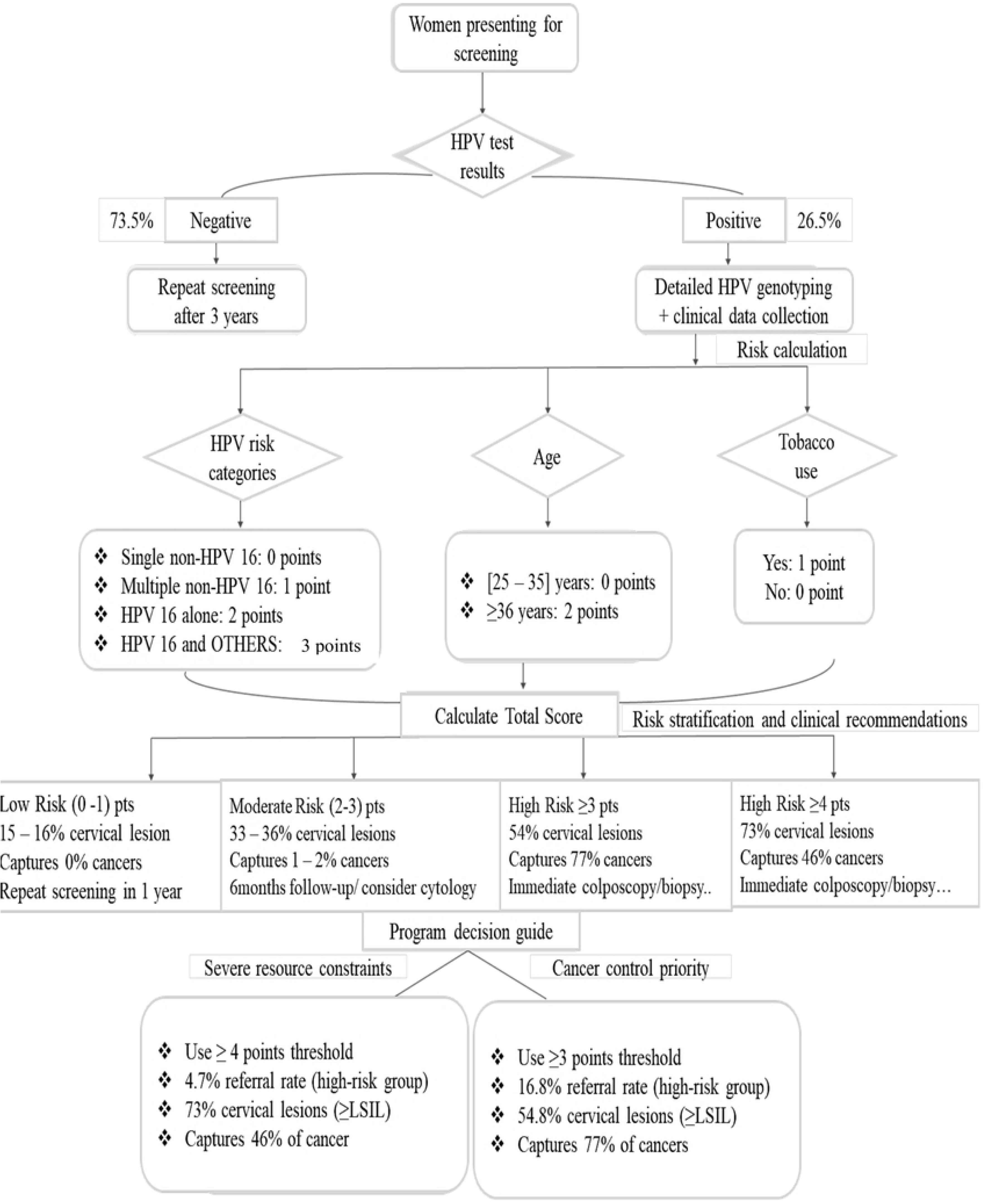
Algorithm for risk-stratified HPV-positive triage. The flowchart illustrates clinical decision pathways based on HPV genotype, age, and tobacco use. The two-threshold design enables flexible adaptation to programmatic priorities (resource efficiency vs. cancer case-finding).

## Discussion

This study was designed within a pragmatic public health framework, where methodological choices were guided by real-world constraints rather than ideal experimental conditions. In low-resource settings, screening strategies must balance accuracy with feasibility, cost, and scalability. We developed and internally validated a genotype-specific risk-stratification model in that light.

### Principal findings and interpretations

The final model confirmed the prevalent oncogenic effect of HPV 16, particularly when present in co-infections (other HR-HPV types), which conferred a markedly elevated risk (OR = 28.7, firth-corrected OR=12.40); all women in this category had an advanced cervical lesion (Table 3). This aligns with findings from numerous studies globally and across sub-Saharan Africa (SSA) that highlight the oncogenic potential of HPV 16 and suggest synergistic effect in multiple infections [11-14]. The hierarchical HPV categorization we employed (moving from single non-16 to HPV 16 with co-infections) successfully translated complex molecular data (HPV) into clinically intelligible and monotonic risk gradient. This gradient is the cornerstone of our scoring system, distinguishing it from current screening tests and aligning with the need for risk-based management in screening programs.

Our model demonstrated moderate but clinically meaningful discriminative ability (AUC = 0.702, 95% CI: 0.64–0.76, p<0.001, Fig 2), which represents a significant advancement in a setting where objective triage tools are largely absent. In cervical cancer triage, the negative predictive value (NPV) is often more critical than AUC for ensuring clinical safety, as it determines the tool’s ability to reliably exclude low-risk women from unnecessary procedures. Our model achieved an excellent NPV exceeding 95% at both implementation thresholds, confirming its safety for deferring intensive follow-up in low-risk women. While this AUC is lower than those reported for machine learning models in research settings (AUC>0.80) [15], it substantially outperforms the visual inspection with acetic acid (VIA) currently used in Cameroon, which suffers from poor reproducibility and subjective interpretation.

Our score offers a significant clinical advantage over VIA by providing a more objective and reliable assessment of a woman’s risk for cervical cancer. While the Negative Predictive Value (NPV) of VIA in Cameroon typically ranges from 70% to 85% [16], our risk score achieves an NPV exceeding 94%. Clinically, this disparity is profound: an 80% NPV implies that 20% of high-risk women will receive “false reassurance” and be missed by the screening, whereas our tool identifies < 6% as false negatives. By replacing the subjective, experience-dependent nature of VIA with a standardized, genotype-informed calculation, we ensure greater reproducibility and a much higher level of safety for women in resource-limited settings.

The Hosmer-Lemeshow test confirmed excellent model calibration (p=0.399), indicating that predicted risk estimates accurately reflect observed event rates across the entire risk spectrum. Beyond this, the calibration plot (Fig 3) confirmed a high level of agreement between predicted probabilities and observed outcomes. The proximity of the data points to the 45-degree diagonal line suggests that the model is well-calibrated across the entire spectrum of HPV-related risk. Again, our calibration analysis showed zero observed lesion proportion in the lower risk strata, closely matching the model’s predictions. Similarly, the model maintained its accuracy in the higher risk strata (predicted risk of 37.7% vs. observed 34.6%). This precision at the higher end of the scale is essential for ensuring that women with the highest likelihood of disease are prioritized for immediate intervention, aligning with the WHO’s ’screen-and-treat’ objectives^2^ while optimizing the use of limited diagnostic resources.

The retention of age (≥36 years) and tobacco use as significant predictors in our final model emphasizes the importance of integrating simple clinical information to enhance risk assessment beyond HPV status alone. Age may reflect cumulative years of exposure to HPV, decreases in immune response oversight of HPV, and the natural history of persistence of HPV. Older women with persistent HPV infection have disproportionately high risks for developing cervical precancerous lesions and cancer [9]. Tobacco increases carcinogenic damage to cervical tissue and affects the persistence of HPV [9].

HIV was not retained in the final model due to sparse-data instability in this specific cohort (9.4% HIV-positive). This does not diminish its clinical importance: women living with HIV in SSA face a 2- to 6-fold higher risk of HPV persistence and cervical cancer progression^11,12^. Larger targeted cohorts enriched for HIV-positive participants are needed to determine whether the scoring thresholds require adjustment in this high-risk subgroup.

### Methodological strengths

We explicitly recognize three methodological choices. First, our iterative bootstrap-guided development strategy (for diagnosing instability) represents a principled, transparent approach to the HPV sparsity problem that is reproducible and biologically motivated. Second, the strict outcome separation between predictor selection (≥LSIL) and final validation (≥HSIL) prevents outcome-dependent overfitting, a common source of optimistic bias. Third, the reporting follows the TRIPOD guideline (Transparent Reporting of a multivariable prediction model for Individual Prognosis Or Diagnosis), ensuring methodological transparency and replicability.

### Clinical and public health implications

The proposed scoring system and the two-threshold implementation algorithm offer a pragmatic solution for limited resource settings. Women in the low-risk group (0 – 1 points) comprise most of the screened population, have a very low probability of significant disease (<17% cervical lesions, 0% cancer) and can be safely managed by repeating screening in one year. Those in the moderate-risk group warrant closer surveillance due to an intermediate profile (33 - 36% cervical lesions, 1 – 2% cancers) and are recommended enhanced monitoring with 6-month follow-up and Pap smear. Women in the high-risk group (≥3 or ≥4 points) require immediate diagnostic evaluation (colposcopy, biopsy). The conservative threshold (≥4 points) is tailored for settings with severe resource constrains, efficiently concentrating efforts on the 4.7% of women at highest risk (PPV = 58%), minimizing referrals. Conversely, the sensitive threshold (≥3 points) is designed for programs aiming to maximize cancer detection, capturing 77% of cancers while referring 16.8% of women.

This flexible approach allows ministries of health to adapt the same core tool to local resource availability and public health priorities, ensuring that limited clinical resources are directed to the women who need them most while safely minimizing intervention for those at low risk. The tool leverages existing laboratory infrastructure (HPV genotyping) and adds minimal data collection burden (age, smoking status), enhancing its feasibility for integration into national screening programs.

### Limitations

- Hospital-based recruitment in pathology and gynecology units enriches the sample for higher-risk women, as evidenced by a ≥HSIL prevalence of 14.6% approximately double the estimated community-level prevalence of 5–8% in Cameroon^11^. Consequently, PPV estimates reported here are likely higher than would be achieved in general community screening contexts, while NPV may be even more reassuring in lower-prevalence settings.
- The cross-sectional nature precludes assessment of the scoring system’s ability to predict future HPV persistence, lesion progression, or treatment response which are the most clinically relevant endpoints for a triage tool.
- Cytology was used as diagnostic reference rather than histology (biopsy), which has higher sensitivity for high grade lesions. Misclassification of true ≥HSIL cases as lower grades on cytology may have attenuated model performance.
- Bootstrap resampling corrects for within-sample optimism but does not replace prospective external validation in geographically independent populations. The scoring system should not be deployed clinically until validated in geographically independent cohorts with different epidemiological profiles.
- Although HIV-positive status was not retained in the final model owing to insufficient discriminatory power at the threshold tested, a trend was observed: women with a positive HIV status had a higher prevalence of ≥HSIL (22%) compared with HIV-negative women (14%). Future work should explore whether adjusting the scoring threshold for HIV-positive women improves triage performance in HIV-endemic populations.

### Conclusion

This risk-stratification model is a proof-of-concept and represents a pragmatic and scalable solution to optimize HPV-based screening in low-resource settings. It generates a clear monotonic risk gradient, maintains NPV >94% at both clinical thresholds, and is designed for flexible adaptation to local resource constraints and programmatic priorities. By reducing unnecessary referrals while maintaining clinical sensitivity, it has the potential to significantly improve the efficiency of cervical cancer prevention programs in sub-Saharan Africa.

## Data Availability

The datasets generated and analyzed during this study are not publicly available, owing to restrictions stipulated in ethical approval to protect participant confidentiality. Anonymized data may be made available from the corresponding author upon reasonable request, subject to approval by the principal investigators and the ethics committee.

## Acknowledgements

We gratefully acknowledge the participation of all women who took part in this study. We thank the directors, clinical, and laboratory staff of the Bafoussam Regional Hospital, Sangmelima Reference Hospital, Garoua Regional Hospital, and Yaoundé Gynecology, Obstetric and Pediatric Hospital. We also acknowledge the laboratory teams at the Chantal Biya International Reference Centre (CIRCB) and the Pathology Laboratory of the Bafoussam Regional Hospital for their collaboration in sample analysis.

## Supporting files

**S1 Table**

**S2 Table**

**S3 Table**

**S1 Fig**

**S1 Note**

